# Bias from small-count suppression in county-level cancer disparity estimates: a calibrated simulation study

**DOI:** 10.64898/2026.06.05.26355021

**Authors:** Kamalakanta Gahan

## Abstract

**Background:** Area-level cancer disparities are routinely estimated from public county data in which rates based on small counts (fewer than 16 cases or deaths) are suppressed. Analysts typically drop suppressed counties (complete-case analysis). Because suppression depends on case counts—tied to population size and demographic composition—this missingness may be informative, but its effect on the disparity estimate has not, to our knowledge, been quantified.

**Methods:** In a cross-sectional ecological study of 3,143 U.S. counties (analytic sample 3,018 with computable exposure) using one frozen public release of NCI State Cancer Profiles incidence and mortality data and ACS 2018–2022 5-year data, we estimated the most-versus least-deprived ICE(race+income) quintile rate ratio (RR) and rate difference for female breast, stomach, and cervix cancers under four suppression-handling methods: complete-case, available-case, bounding, and model-based small-area estimation. We characterized which counties were erased, and—following the ADEMP framework—ran a Monte Carlo simulation (1,000 replicates per cell; Monte Carlo standard error of bias approx. 0.0025) calibrated to the release to measure bias against a known truth. Analyses were pre-registered.

**Results:** The suppressed fraction rose with rarity: 7.4% of counties for breast, 61.3% for stomach, and 75.7% for cervix incidence. Suppression was concentrated in the most-deprived quintile (cervix, 81.8% suppressed vs 63.8% least-deprived) and overwhelmingly removed rural rather than minority residents (cervix: 81% of the rural but 9% of the minority population erased). For breast (little suppression) the RR was 0.87 (95% CI 0.85–0.89) and identical across methods; for cervix incidence the complete-case RR (1.56) exceeded the model-based estimate (1.50), and for cervix mortality (91% suppressed) complete-case (1.86) exceeded model-based (1.56) by 16%, with an assumption-free bounding interval so wide (0.81–6.05) that the disparity was effectively unidentified. In calibrated simulation, population-weighted complete-case bias was small (<2%) at the observed deprivation–county-size correlation and grew with rarity, threshold, and unweighted aggregation; its direction was conditional, becoming positive (over-estimation) as deprived counties became smaller.

**Conclusions:** Complete-case handling of suppressed counties over-estimates rare-cancer area disparities relative to methods that retain them, while silently erasing most of the rural and most-deprived communities the estimate is meant to represent. The effect is negligible for common cancers and grows with rarity. Public-data disparity analyses should report the suppressed fraction and use bounded or model-based estimates by default.

## Introduction

Structural determinants of health, and residential segregation in particular, are central drivers of cancer inequities in the United States. The Index of Concentration at the Extremes for race and income, ICE(race+income), is a validated measure of racialized economic segregation that jointly captures the spatial concentration of privilege and deprivation, and it has been applied to cancer outcomes to reveal inequities that single-axis measures of poverty or race miss (Krieger et al., 2016, *Cancer Causes Control*; Larrabee Sonderlund et al., 2022). Area-level disparity estimates built on such measures increasingly inform cancer-control priorities and resource allocation.

In practice, these analyses rest on public, county-level registry data—the National Cancer Institute’s State Cancer Profiles and similar products—because the restricted-access microdata that would permit individual-level analysis require a data-use agreement. Public county data, however, carry a structural feature that is rarely treated as part of the analysis: small-count suppression. To protect confidentiality and avoid reporting unstable statistics, rates based on fewer than 16 cases or deaths in the reporting period are withheld (NCI State Cancer Profiles; CDC/NPCR–United States Cancer Statistics technical documentation), and in bulk data these strata are simply absent. Such omissions fall hardest on small and rural areas, exactly where prior work shows the cancer burden and its disparities are most easily missed (Houston et al., 2018), and where rare, low-count cancers in very small areas demand special statistical handling (Etxeberria et al., 2022).

Analysts routinely accommodate suppression by dropping the affected counties—a complete-case analysis— and proceeding as though the remaining counties were the population of interest. Yet how much, and in which direction, this practice distorts the area-level disparity estimate has not, to our knowledge, been quantified, and whether the resulting erasure of counties is itself patterned by deprivation has not been examined. Because suppression is governed by the case count, which is tied to population size and demographic composition, there is reason to expect the missingness to be informative rather than benign.

We quantify, using a calibrated simulation and a single public release of county cancer data, how small-count suppression biases county-level ICE cancer-disparity estimates and which communities it erases.

## Methods

We report the observational analysis following the STROBE guideline for cross-sectional studies and the simulation study following the ADEMP framework (aims, data-generating mechanisms, estimands, methods, performance measures) of Morris, White, and Crowther (Morris et al., 2019).

### 1. Study design and data sources

This was a cross-sectional ecological analysis of United States counties complemented by a Monte Carlo simulation. County-level age-adjusted cancer incidence and mortality rates (“latest 5-year average,” per 100,000) were obtained from the National Cancer Institute’s State Cancer Profiles via a public, version-tagged bulk mirror of the resource (release tag 2026-06-01). Area socioeconomic data came from the American Community Survey (ACS) 2018–2022 5-year estimates (U.S. Census Bureau). Rurality used the Rural/Urban designation distributed with the release, with USDA Rural-Urban Continuum Codes (RUCC) 2023 reserved for sensitivity analysis. All data are publicly available, aggregated to the county level, and de-identified.

### 2. Unit of analysis and study population

#### The unit of analysis is the county (or county-equivalent), not the individual

The study population comprised the 3,143 counties of the 50 states and the District of Columbia; Puerto Rico and other territories (state FIPS 72, 78, 60, 66, 69) were excluded. Of these, 3,018 counties had a computable ICE(race+income) value (the 125 excluded lacked the required ACS household-income tables) and constituted the analytic sample; counts by exposure quintile are reported in Results (STROBE participant flow).

### 3. Exposure — ICE(race+income)

The exposure was the Index of Concentration at the Extremes for racialized economic segregation (Krieger et al., 2016, *Cancer Causes Control*; Krieger et al., 2016, *Am J Public Health*). For county *i*, ICE = (A - P) / T, where A = White non-Hispanic households with income >= $100,000 (ACS table B19001H, income brackets 014–017), P = households of color (all households minus White non-Hispanic) with income < $25,000 (B19001 brackets 002–005 minus the corresponding B19001H brackets, floored at zero), and T = total households (B19001_001). ICE ranges from −1 (all low-income households of color) to +1 (all high-income White households); **lower values denote greater deprivation**. Counties were assigned to population-weighted quintiles (each approx. 20% of the total population, weighting by ACS county population [B01003]); quintile cut points were computed on the full county universe before any outcome was examined. Quintiles are labelled Q1 = most deprived (lowest ICE) to Q5 = least deprived.

### 4. Outcome and the suppression rule

Outcomes were county age-adjusted incidence (primary) and mortality (parallel) rates per 100,000, in the all-races, all-stages, all-ages stratum (sex female for breast and cervix; both sexes for stomach). Per State Cancer Profiles and CDC/NPCR–United States Cancer Statistics documentation, **a rate is suppressed when the underlying count for the reporting period is fewer than 16**, applied by the data producer for statistical stability and confidentiality; in the bulk data, suppressed strata are absent rows rather than flagged values. We verified this empirically: the minimum non-suppressed average annual count was approx. 3 (approx. 16 over the 5-year period), consistent with the documented rule. The exact threshold is treated as load-bearing and is varied in sensitivity analysis (§9).

### 5. Estimand

**The estimand is the ratio (rate ratio, RR) and the difference (rate difference, RD) of age-adjusted rates between the most-deprived (Q1) and least-deprived (Q5) ICE quintiles**, with a population-weighted gradient across all five quintiles as a secondary summary. Each estimand is defined two ways: **population-weighted** (quintile rate = population-weighted mean of county rates; primary) and **unweighted county-mean** (secondary), because the latter is common in practice and is differentially sensitive to suppression. Naming the estimand independently of the estimator makes explicit what each suppression-handling method targets.

### 6. Missing-data framework and suppression-handling methods

Because suppression is determined by the case count—itself a function of population size and demographic composition—the missingness is **informative / missing-not-at-random (MNAR)**, and complete-case analysis is therefore non-ignorable. We compared four handlings and state the assumption of each: (1) **complete-case** (analyze counties with a reported rate; the de facto standard under critique); (2) **available-case** (the complete-case estimate reported alongside explicit per-quintile retention of counties and population); (3) **bounding** (assumption-free Manski-style worst/best-case bounds: each suppressed county’s true rate lies between 0 and (threshold - 1) cases per person-time, and the extremal rate ratio and rate difference are obtained by imputing the most- and least-deprived quintiles at opposite extremes of this range); (4) **model-based small-area estimation** that borrows strength across counties, in the spirit of hierarchical Bayesian small-area smoothing of sparse county rates (Schootman et al., 2010) and multilevel count models for ICE (Feldman et al., 2019). The executed reference implementation used an empirical-Bayes state-prior shrinkage estimator (each suppressed county imputed by its state’s population-weighted mean rate, national fallback) as a transparent stand-in for a full hierarchical Poisson model. A >=10% relative difference between complete-case and the model-based (or bounding-midpoint) estimate was pre-specified as non-trivial. General principles of bias under informative missingness follow Sterne et al. (2009).

### 7. Simulation study (reported under ADEMP)

**Aims**. To quantify the direction and magnitude of suppression-induced bias in the disparity estimand (§5) relative to a known ground truth. **Data-generating mechanisms**. For each replicate, N = 3,143 counties were generated. County log-population was drawn from a normal distribution (mean 10.272, SD 1.50) calibrated to the empirical U.S. county size distribution, correlated with a standard-normal latent deprivation score through a tunable correlation. Counties were assigned to deprivation quintiles. The true county rate was log-linear in deprivation, rate = base x exp(beta.z), with beta the log rate ratio per SD of deprivation. Expected period counts were rate x population x 5 person-years; observed counts were Poisson (and, in sensitivity, gamma-Poisson overdispersed) draws. Counties with a simulated period count below the suppression threshold were dropped, mirroring the empirical mechanism. **Estimands**. Identical to §5 (population-weighted and unweighted Q1-vs-Q5 RR and RD). **Methods**. The suppression-handling approaches of §6 (primary comparison: complete-case vs known truth). **Performance measures**. Bias (estimate - truth), percent (relative) bias, and the Monte Carlo standard error (MCSE) of bias. **nSim = 1**,**000 replicates per scenario cell**; this was justified by the target MCSE: with an observed replicate-level SD of the RR bias of approx. 0.079, ~63 replicates suffice to keep MCSE < 0.01 on the rate-ratio scale, so 1,000 gives MCSE approx. 0.0025 (about 0.06% of the true ratio). The fully crossed grid varied base incidence (5, 15, 50, 150 per 100,000), the deprivation–rate association (RR per SD 1.0, 1.3, 1.7), the deprivation–county-size correlation (0, −0.3, −0.6), and the suppression threshold (period count 10, 16, 25). All random draws used reproducible seeds (base seed 20260604, offset by scenario index).

### 8. Calibration

Structural simulation parameters were fit to the analyzed release rather than assumed. The county size distribution (log-mean 10.272, log-SD 1.50) and the deprivation–county-size correlation (−0.30; median population 146,684 in Q5 versus 18,580 in Q1, ratio 0.24) were estimated from ACS county population and ICE deprivation. The overdispersion parameter was approximated from the spread of reported county rates and treated as a sensitivity input rather than a point estimate. The simulated suppression rate by deprivation quintile was checked against the empirical suppression-by-quintile table (PROVENANCE.md).

### 9. Pre-specified sensitivity analyses

Pre-registered before analysis (OSF; https://doi.org/10.17605/OSF.IO/BJZR5): alternative suppression thresholds (10, 25 in addition to 16); an alternative self-contained deprivation index and the CDC/ATSDR SVI as alternative exposures; population-weighted versus unweighted aggregation; and variation of the overdispersion parameter. The empirical analysis was also run for mortality in parallel to incidence.

### 10. Software and reproducibility

Analyses are scripted, seeded, and version-pinned. The executed reference implementation is Python 3.10 with DuckDB (duckdb 1.5.3, pandas 2.3.3, numpy 2.2.6, matplotlib 3.10.8); a canonical port of the estimator and the four suppression-handling methods is provided in R. Code, derived data, the provenance log, and figure scripts are public (https://github.com/kgahanduke-25/suppression-bias); the study was pre-registered (https://doi.org/10.17605/OSF.IO/BJZR5). The release tag (2026-06-01) and ACS vintage (2018–2022 5-year) are frozen.

### 11. Ethics

The study is a secondary analysis of publicly available, de-identified, aggregate data and does not constitute human-subjects research; it was therefore exempt from institutional review board review.

## Results

### Analytic sample

Of the 3,143 U.S. counties, 3,018 had a computable ICE(race+income) value and were included; the remainder lacked ACS household-income tables. Counties were strongly patterned across ICE quintiles (Table 1). Median county population rose from 18,890 (IQR 9,483–41,986) in the most-deprived quintile (Q1) to 53,381 (IQR 19,794–167,567) in the least-deprived (Q5); the share urban rose from 26.6% to 71.1%; mean percent Black fell from 26.9% (SD 22.2) to 3.2% (SD 4.1); and median household income rose from $51,424 to $89,975 (standardized difference Q1 vs Q5 = 2.79). Reported breast-cancer incidence was higher in less-deprived counties (median 123.1 vs 137.8 per 100,000, Q1 vs Q5), whereas reported cervix incidence was higher in more-deprived counties (9.9 vs 6.0 per 100,000).

**Table 1.**
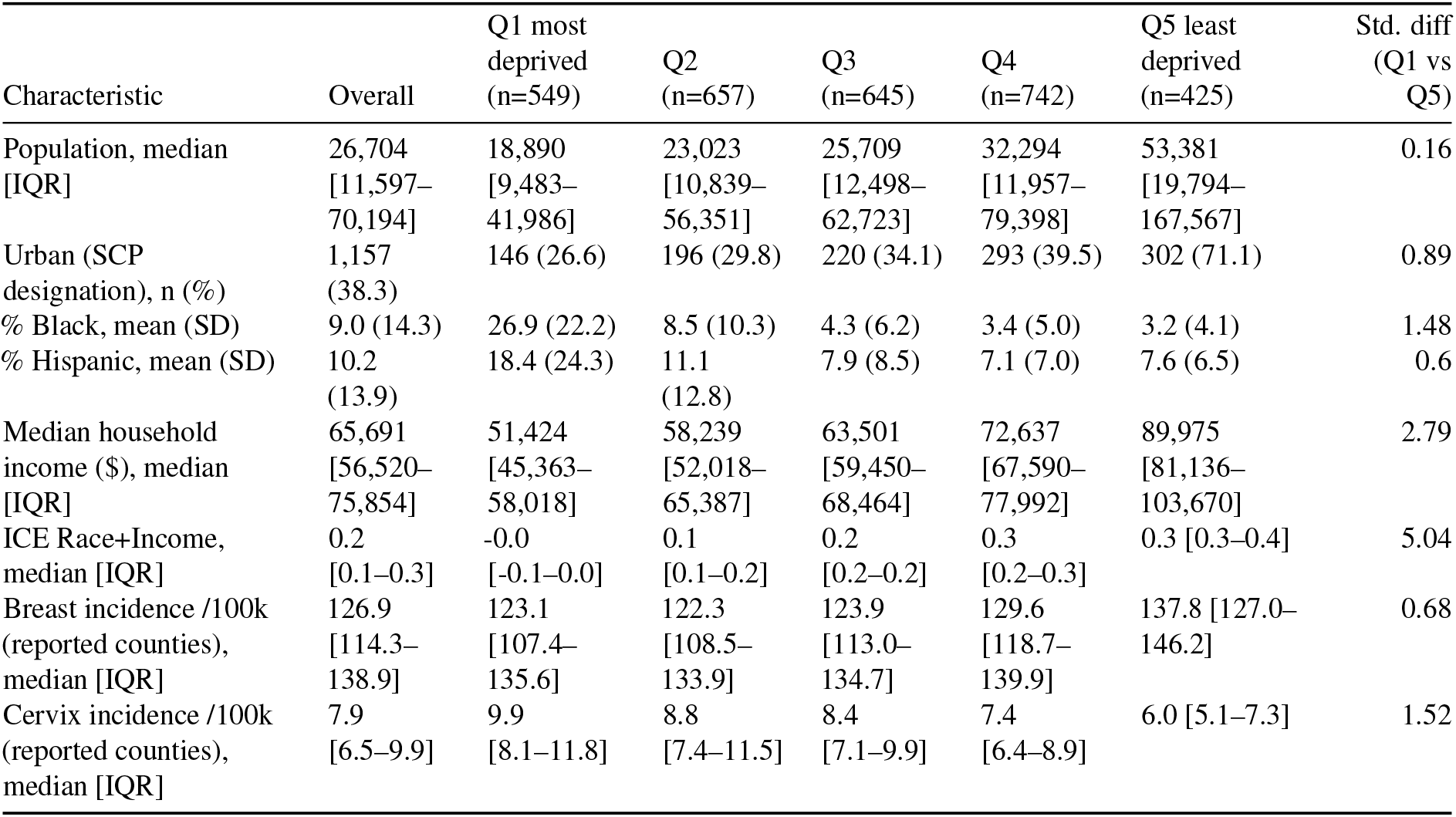
Characteristics of US counties (n=3,143; analytic n=3,018) by ICE Race+Income quintile, ACS 2018–2022. Unit = county; population-weighted ICE quintiles (Q1 = most deprived). Continuous: median [IQR] or mean (SD) as labelled; categorical: n (%). Std. diff = standardized difference (Q1 vs Q5). Quintiles are population-weighted, so county counts differ across quintiles.

| Characteristic | Overall | Q1 most deprived (n=549) | Q2 (n=657) | Q3 (n=645) | Q4 (n=742) | Q5 least deprived (n=425) | Std. diff (Q1 vs Q5) |
| --- | --- | --- | --- | --- | --- | --- | --- |
| Population, median [IQR] | 26,704 [11,597–70,194] | 18,890 [9,483–41,986] | 23,023 [10,839–56,351] | 25,709 [12,498–62,723] | 32,294 [11,957–79,398] | 53,381 [19,794–167,567] | 0.16 |
| Urban (SCP designation), n (%) | 1,157 (38.3) | 146 (26.6) | 196 (29.8) | 220 (34.1) | 293 (39.5) | 302 (71.1) | 0.89 |
| % Black, mean (SD) | 9.0 (14.3) | 26.9 (22.2) | 8.5 (10.3) | 4.3 (6.2) | 3.4 (5.0) | 3.2 (4.1) | 1.48 |
| % Hispanic, mean (SD) | 10.2 (13.9) | 18.4 (24.3) | 11.1 (12.8) | 7.9 (8.5) | 7.1 (7.0) | 7.6 (6.5) | 0.6 |
| Median household income (\$), median [IQR] | 65,691 [56,520–75,854] | 51,424 [45,363–58,018] | 58,239 [52,018–65,387] | 63,501 [59,450–68,464] | 72,637 [67,590–77,992] | 89,975 [81,136–103,670] | 2.79 |
| ICE Race+Income, median [IQR] | 0.2 [0.1–0.3] | -0.0 [-0.1–0.0] | 0.1 [0.1–0.2] | 0.2 [0.2–0.2] | 0.3 [0.2–0.3] | 0.3 [0.3–0.4] | 5.04 |
| Breast incidence /100k (reported counties), median [IQR] | 126.9 [114.3–138.9] | 123.1 [107.4–135.6] | 122.3 [108.5–133.9] | 123.9 [113.0–134.7] | 129.6 [118.7–139.9] | 137.8 [127.0–146.2] | 0.68 |
| Cervix incidence /100k (reported counties), median [IQR] | 7.9 [6.5–9.9] | 9.9 [8.1–11.8] | 8.8 [7.4–11.5] | 8.4 [7.1–9.9] | 7.4 [6.4–8.9] | 6.0 [5.1–7.3] | 1.52 |

### Extent and patterning of suppression (Aim 2)

The fraction of counties suppressed under complete-case analysis increased with site rarity: 7.4% for breast, 61.3% for stomach, and 75.7% for cervix incidence (Table 3). Suppression was greater in more-deprived quintiles: for cervix, 81.8% of Q1 (most-deprived) counties were suppressed versus 63.8% of Q5 (Table 3B).

The population erased was concentrated among rural and small-county residents rather than minority residents (Figure 2). For cervix incidence, complete-case analysis removed 17.4% of the total population, 81.4% of the rural population, but only 9.3% of the minority population; corresponding figures were 9.7%, 52.8%, and 5.1% for stomach, and 0.2%, 1.3%, and 0.1% for breast. Within the most-deprived cervix quintile, 78.3% of the rural population but 11.9% of the minority population was erased (Table 3B).

### Disparity estimates by suppression-handling method (Aim 1)

For breast-cancer incidence (7.4% suppressed; 90% of most-deprived counties retained), the most-versus least-deprived rate ratio was 0.87 (95% CI 0.85–0.89) and was unchanged across all handling methods— population-weighted complete-case 0.87, unweighted 0.90, bounding interval 0.87–0.87, model-based 0.87 (Table 2A; Figure 1).

**Table 2A.** Estimated breast-cancer incidence disparity (most- vs least-deprived ICE quintile) by suppression-handling method.

| Handling method | Rate ratio (95% CI) | Rate difference (/100k) | Counties included | % most-deprived counties retained |
| --- | --- | --- | --- | --- |
| Complete-case (population-weighted) | 0.87<br>(0.85–0.89) | -18.4 | 2,794 | 90 |
| Complete-case (unweighted county-mean) | 0.90<br>(0.88–0.92) | -13.8 | 2,794 | 90 |
| Available-case (= complete-case; retention shown) | 0.87<br>(0.85–0.89) | -18.4 | 2,794 of 3,018 | 90 |
| Bounding — impute 0 | 0.87 | -18.8 | 3,018 | 100 |
| Bounding — impute threshold | 0.87 | -18.4 | 3,018 | 100 |
| Model-based (small-area) | 0.87<br>(0.85–0.89) | -18.4 | 3,018 | 100 |

**Table 2B.** Estimated cervix-cancer incidence disparity by suppression-handling method.

| Handling method | Rate ratio (95% CI) | Rate difference (/100k) | Counties included | % most-deprived counties retained |
| --- | --- | --- | --- | --- |
| Complete-case (population-weighted) | 1.56<br>(1.46–1.67) | 3.3 | 732 | 18 |
| Complete-case (unweighted county-mean) | 1.61<br>(1.49–1.73) | 3.9 | 732 | 18 |
| Available-case (= complete-case; retention shown) | 1.56<br>(1.46–1.67) | 3.3 | 732 of 3,018 | 18 |
| Bounding — impute 0 | 1.24 | 1.5 | 3,018 | 100 |
| Bounding — impute threshold | 2.03 | 5.2 | 3,018 | 100 |
| Model-based (small-area) | 1.50<br>(1.42–1.59) | 3 | 3,018 | 100 |

**Table 3A.**
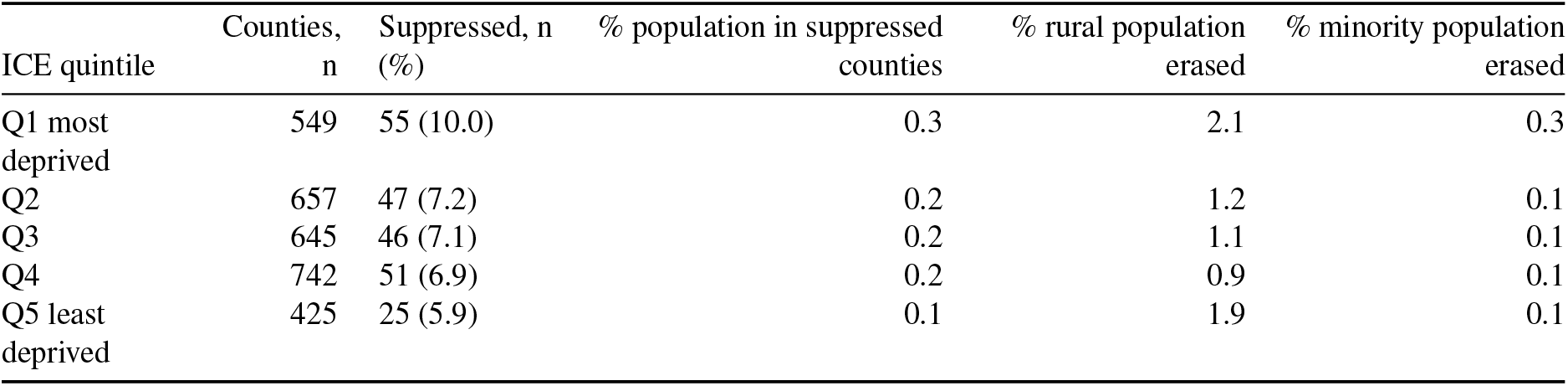
Suppression diagnostics by ICE quintile — breast incidence.

**Table 3B.** Suppression diagnostics by ICE quintile — cervix incidence.

| ICE quintile | Counties, n | Suppressed, n (%) | % population in suppressed counties | % rural population erased | % minority population erased |
| --- | --- | --- | --- | --- | --- |
| Q1 most deprived | 549 | 449 (81.8) | 15 | 78.3 | 11.9 |
| Q2 | 657 | 505 (76.9) | 14.5 | 80.9 | 7 |
| Q3 | 645 | 502 (77.8) | 19.9 | 82.4 | 8.6 |
| Q4 | 742 | 559 (75.3) | 22.9 | 81.1 | 10.4 |
| Q5 least deprived | 425 | 271 (63.8) | 14.7 | 88.7 | 8 |

*Abbreviations:* ICE, Index of Concentration at the Extremes; RR, rate ratio; RD, rate difference; CI, confidence interval; IQR, interquartile range; SCP, State Cancer Profiles. *Source:* NCI State Cancer Profiles release 2026-06-01; ACS 2018–2022 5-year. *Suppression rule:* rates with period count <16 are not reported.

**Figure 1:**
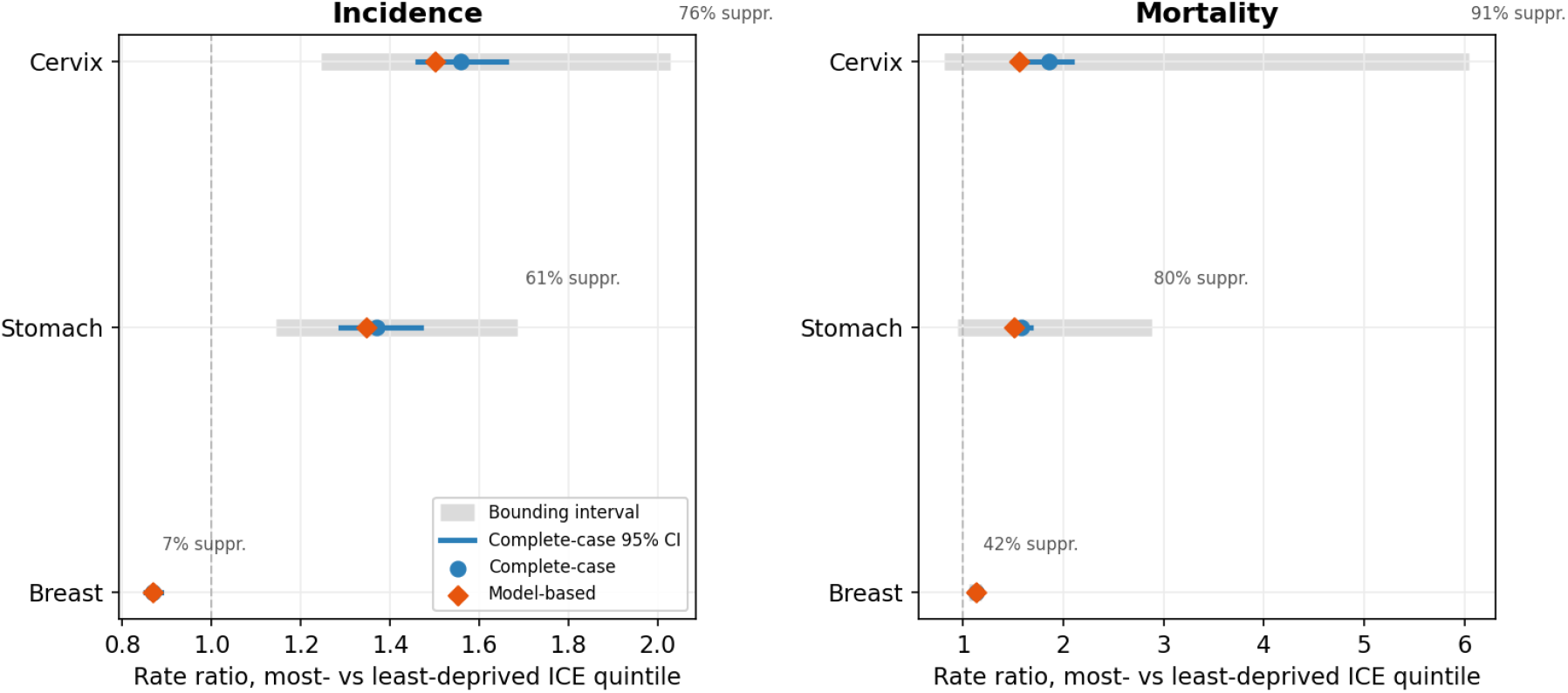
Estimated most- vs least-deprived ICE(race+income) quintile rate ratio (RR) for breast, stomach, and cervix cancer, by suppression-handling method, for incidence (left) and mortality (right). Unit = county; blue circle = population-weighted complete-case point estimate with 95% bootstrap CI; orange diamond = model-based small-area estimate; grey bar = bounding interval; dashed line = no disparity (RR=1); labels =% of counties suppressed. Source: NCI State Cancer Profiles release 2026-06-01; ACS 2018–2022.

**Figure 2:**
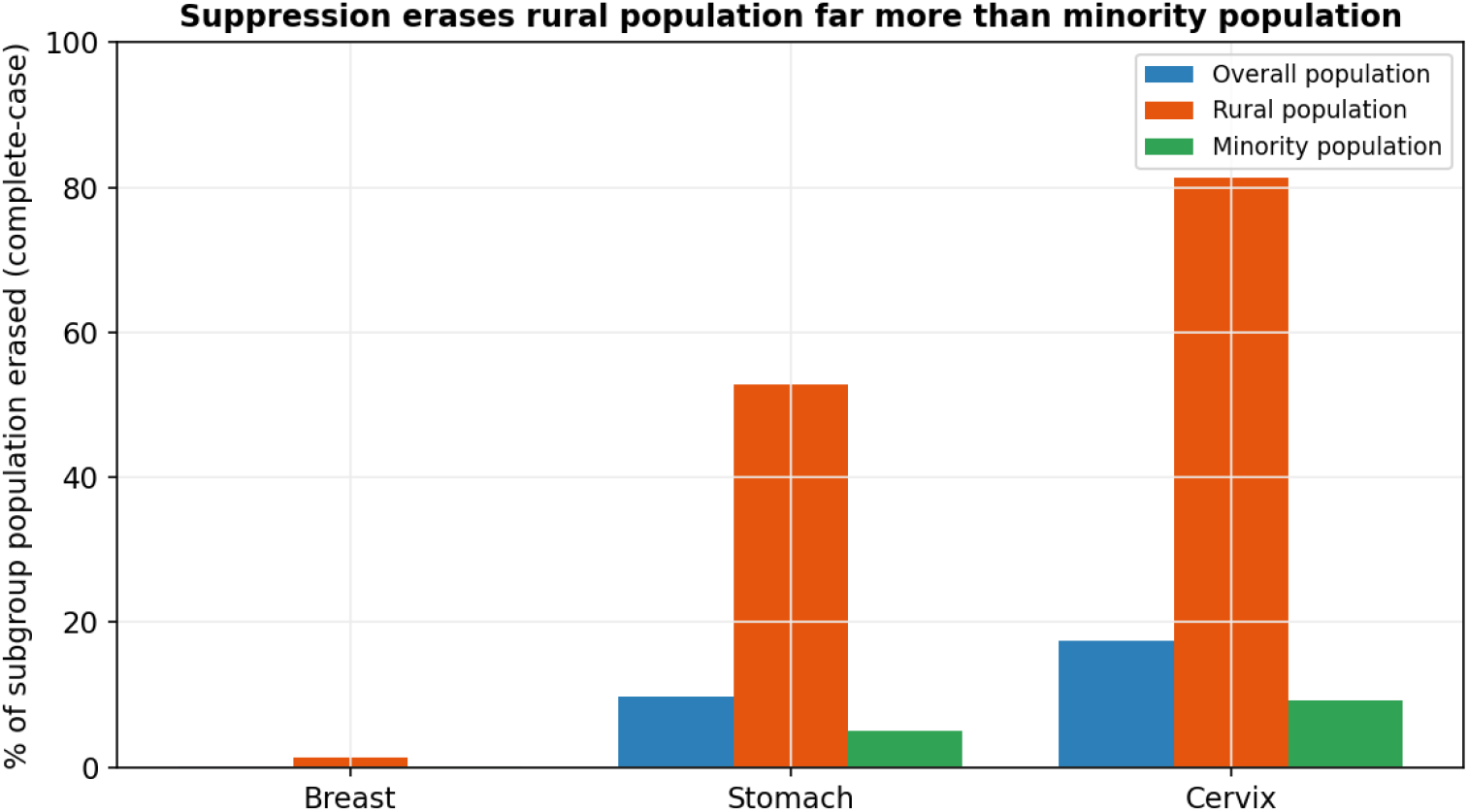
Percent of each subgroup’s population erased under complete-case analysis (suppressed counties), by site. Unit = county; subgroups are overall, rural, and minority (non-White) population. Source: NCI State Cancer Profiles release 2026-06-01; ACS 2018–2022.

For cervix-cancer incidence (75.7% suppressed; only 18% of most-deprived counties retained), the population-weighted complete-case rate ratio was 1.56 (95% CI 1.46–1.67); the unweighted estimate was 1.61 (1.49– 1.73); the assumption-free bounding interval spanned 1.24–2.03; and the model-based estimate was 1.50 (1.42–1.59), a 3.6% relative reduction from complete-case (Table 2B). Divergence between methods was largest for the most heavily suppressed outcome, cervix mortality (90.7% suppressed), where the complete-case rate ratio was 1.86 (95% CI 1.65–2.09), the model-based estimate 1.56 (a 16.0% relative reduction), and the assumption-free bounding interval was so wide (0.81–6.05) that the disparity was effectively unidentified (results/aim1_disparities.csv). Across sites, the relative difference between complete-case and model-based estimates rose with the suppression fraction: ~0% (breast incidence, 7.4%), −3.6% (cervix incidence, 75.7%), −4.6% (stomach mortality, 80.4%), and −16.0% (cervix mortality, 90.7%) (Figure 1).

### Simulation against known truth (Aim 3)

In simulation, the direction and magnitude of complete-case bias depended on the data-generating process (Figure 3). At the empirically calibrated deprivation–county-size correlation (−0.30), the population-weighted complete-case rate ratio was within 2% of the true rate ratio across all base incidences, including at ~70% suppression; unweighted county-unit estimates were biased approximately twice as much (results/aim3_calibrated_sensitivity.csv). The relative bias became positive (over-estimation, up to +6%) only when the deprivation–size correlation was steeper than observed (−0.6) and the suppression threshold was high; when county size was independent of deprivation (correlation 0) the bias was slightly negative. In all calibrated scenarios the magnitude of the population-weighted bias was small relative to the share of counties and rural population erased.

**Figure 3:**
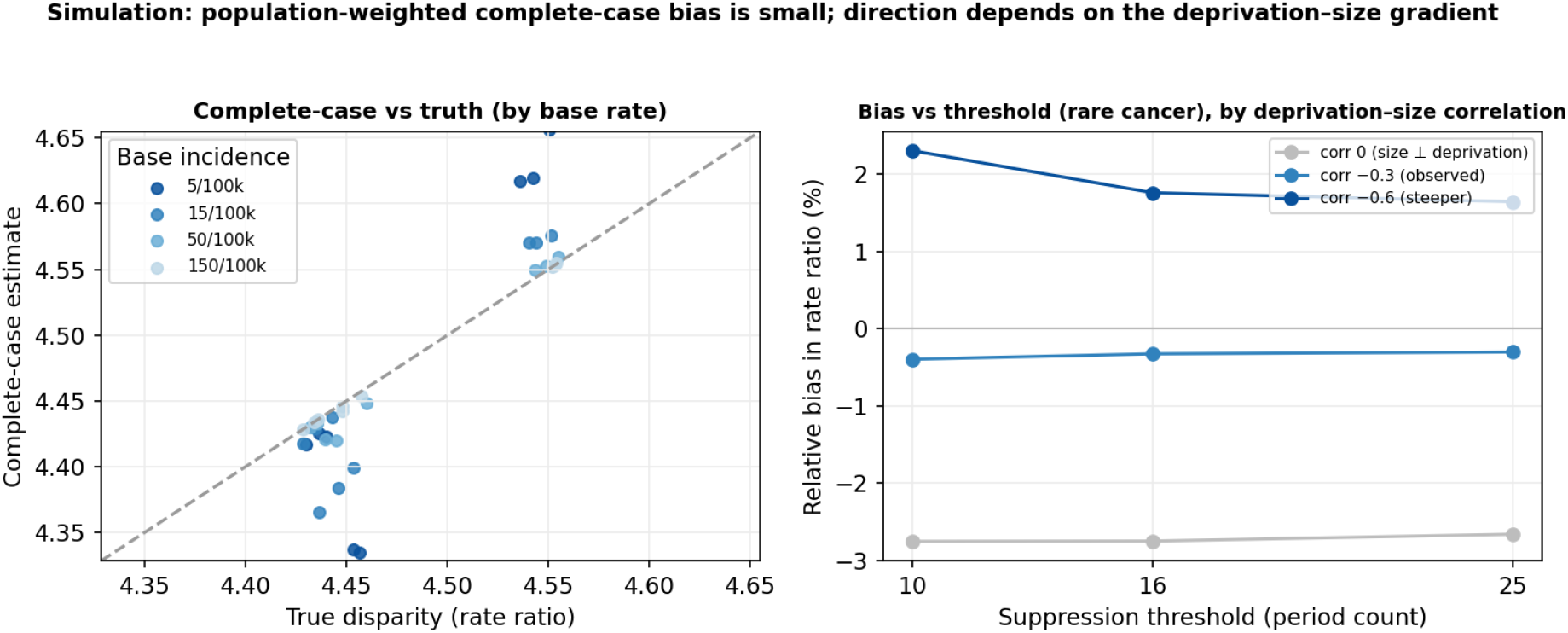
Simulation against known truth. Left: complete-case rate-ratio estimate versus the true rate ratio across scenarios, by base incidence (identity line dashed). Right: relative bias in the rate ratio versus suppression threshold for a rare cancer, by the deprivation–county-size correlation (observed = −0.30). Population-weighted estimand; 1,000 replicates per cell (Monte Carlo SE of bias approx. 0.0025).

## Discussion

### Principal findings

Handling suppressed small-count counties by the de facto standard—complete-case analysis—distorts area-level cancer disparity estimates, and it does so in a direction and to a degree that depend on cancer rarity. For a common cancer (female breast), suppression removed few counties and the most-versus least-deprived ICE disparity was unchanged across every handling method. For rarer cancers it removed most of the relevant counties—about three-quarters of counties and the large majority of the most-deprived counties for cervix—and complete-case estimates sat above model-based small-area estimates by an amount that grew with the suppressed fraction, reaching a 16% higher rate ratio for the most heavily suppressed outcome (cervix mortality, 91% suppressed). Unweighted county-unit analyses were more affected than population-weighted ones. The single most consistent consequence was not a large shift in the headline ratio but the silent erasure of the very communities the estimate is meant to represent: most of the rural population of high-deprivation areas disappeared from the analyzable data for rare cancers.

### Interpretation in relation to prior work

These results extend a known concern—that national or aggregate estimates understate the burden borne by small and rural areas (Houston et al., 2018)—to the disparity estimate itself, which to our knowledge has not previously been quantified as a function of suppression. The exposure we used, ICE(race+income), is an established and validated measure of racialized economic segregation (Krieger et al., 2016, *Am J Public Health*; Larrabee Sonderlund et al., 2022), and its application to cancer outcomes is precedented (Krieger et al., 2016, *Cancer Causes Control*). Two distinct lines of evidence should be read separately. The simulation provides **internal validity**: against a known truth, it shows that population-weighted complete-case bias is small at the empirically observed deprivation–county-size relationship and that its sign is conditional. The empirical analysis provides **external relevance**: it shows the magnitudes that arise in the actual published data, where complete-case and model-based estimates can diverge materially for rare, heavily suppressed outcomes. The two are complementary, not interchangeable.

### Mechanism

The pattern follows from informative (MNAR) missingness. Suppression deletes counties with few cases, and few cases occur disproportionately in small, rural, and more-deprived counties—precisely the most-deprived end of the ICE distribution, where median county population was roughly one-quarter that of the least-deprived quintile. Within a quintile, removing the lowest-count counties is a non-random thinning, so the surviving most-deprived counties are systematically the larger ones; the more a site’s counts are sparse (rarer cancers, higher thresholds), the more severe and selective the thinning, and the more complete-case estimates drift relative to estimates that retain the suppressed counties. Aggregation matters because population weighting down-weights the small erased counties, partially buffering the point estimate, whereas unweighted county-mean analysis does not.

### Strengths

The study uses only public, aggregate data and is fully reproducible without a data-use agreement; code, derived data, and a provenance log are openly available and the analysis was pre-registered. It pairs a calibrated simulation—whose structural parameters were fit to the same release rather than assumed—with the corresponding empirical analysis, and it states the estimand explicitly and separately from the estimators. Reporting follows STROBE and, for the simulation, the ADEMP framework with Monte Carlo standard errors (Morris et al., 2019).

### Limitations

Several limitations qualify these findings. First, the design is ecological; ICE quintiles describe area context, and no individual-level inference is warranted. Second, suppression is MNAR; the bounding analysis brackets the disparity without distributional assumptions but cannot fully resolve it, and the model-based estimator imposes a borrowing-strength structure whose own assumptions (here, shrinkage toward the state mean) may themselves bias the estimate—our model-based values should be read as one plausible reconstruction, not ground truth. Third, the modifiable areal unit problem applies; counties are a fixed but arbitrary unit and quintile cut points are a modeling choice, which we varied in sensitivity analysis. Fourth, area covariates derived from survey sources (e.g., ACS, and BRFSS-derived measures where used) are themselves model-based estimates carrying sampling and modeling error. Fifth, the overdispersion parameter in the simulation was calibrated approximately and is reported as a sensitivity input. Sixth, the analysis is cross-sectional and based on a single data release; behavior across successive releases is the planned sequel. Finally, we are explicit that for common cancers the absolute bias is modest—suppression does not materially distort the breast-cancer disparity—and we do not claim otherwise.

### Implications and what next

Two practical recommendations follow. When estimating area-level cancer disparities from public county data, investigators should report model-based or bounded estimates rather than complete-case estimates alone, and should always report the suppressed fraction (and which communities it removes) as a standard diagnostic, because a population-weighted point estimate can look stable while most rural and deprived counties have been dropped. Three next steps are concrete: a longitudinal extension tracking how disparity estimates move as suppression patterns change across monthly releases; validation of the suppression-handling methods against restricted-access SEER microdata, where the true small-count rates are observable; and assessment across additional rare sites to map where the empirical bias becomes decision-relevant.

## Conclusion

Small-count suppression handled by complete-case analysis over-estimates rare-cancer area disparities relative to methods that retain suppressed counties, while silently erasing most of the rural and most-deprived communities the estimate is meant to describe. The effect is negligible for common cancers and grows with rarity and with unweighted aggregation. Public-data disparity analyses should report the suppressed fraction and use bounded or model-based estimates as the default.

## Data Availability

All data are publicly available. County cancer statistics were obtained from the NCI State Cancer Profiles public release (seandavi/state-cancer-profile-scraper, tag 2026-06-01); population and ICE covariates from the American Community Survey (2018-2022 5-year, tables B19001, B19001H, and B01003); rurality from USDA Rural-Urban Continuum Codes (2023). All analysis and simulation code, and the pre-registration, are openly available at https://github.com/kgahanduke-25/suppression-bias and https://doi.org/10.17605/OSF.IO/BJZR5. All inputs are public and de-identified; no restricted or identifiable data were used.

## Title page and statements

### Author contributions (CRediT)

K.G.: Conceptualization; Methodology; Software; Formal analysis; Investigation; Data curation; Validation; Visualization; Writing – original draft; Writing – review & editing.

### Ethics / IRB

This study is a secondary analysis of publicly available, de-identified, aggregate data and does not constitute human-subjects research; it was therefore exempt from institutional review board review.

### Data and code availability

All inputs are public and de-identified; no restricted or identifiable data were used. - **Cancer data:** NCI State Cancer Profiles (a joint NCI/CDC resource), obtained from the public scraped release `seandavi/state-cancer-profile-scraper`, **release tag 2026-06-01** (county-level incidence and mortality). - **Covariates:** U.S. Census Bureau American Community Survey **2018–2022 5-year** estimates, tables **B19001, B19001H** (household income, overall and White non-Hispanic) and **B01003** (total population) for ICE; USDA **Rural-Urban Continuum Codes 2023** for rurality. - **Code:** analysis, simulation, and figure scripts are openly available at https://github.com/kgahanduke-25/suppression-bias (commit/tag to cite at submission). Executed reference: Python 3.10 (duckdb 1.5.3, pandas 2.3.3, numpy 2.2.6, matplotlib 3.10.8); canonical R port included. - **Pre-registration:** OSF, https://doi.org/10.17605/OSF.IO/BJZR5. A provenance log (`PROVENANCE.md`) records the exact run order, seeds, and calibrated parameters.

## Acknowledgments

The author thanks the National Cancer Institute and CDC for the State Cancer Profiles resource, the U.S. Census Bureau for the American Community Survey, and the maintainer of the public data release used here.

## Reporting guidelines

The observational analysis is reported in accordance with STROBE (cross-sectional studies); the simulation study is reported following the ADEMP framework (Morris et al., 2019). A completed STROBE checklist is provided as a supplement.

## Data-source documentation (non-journal)

National Cancer Institute, State Cancer Profiles (suppression of rates with counts <16); CDC/NPCR United States Cancer Statistics technical notes (suppression of rates and counts); U.S. Census Bureau, American Community Survey 2018–2022 5-year; USDA Economic Research Service, Rural-Urban Continuum Codes 2023.

