## Supplementary Material (Tables S1-S3) for "Bias from small-count suppression in county-level cancer disparity estimates: a calibrated simulation study"

### Supplementary Table S1. Full simulation grid (108 scenarios)

Population-weighted complete-case rate ratio versus known truth. beta = log rate ratio per SD of deprivation; corr = deprivation-county-size correlation. 1,000 replicates/cell.

| base_rate | beta_logRR_perSD | corr_pop_dep | threshold | mean_suppressed | RR_true | RR_cc | RR_relbias |
|---|---|---|---|---|---|---|---|
| 5 | 0 | 0 | 10 | 0.571 | 1 | 1 | -0.0002 |
| 5 | 0 | 0 | 16 | 0.692 | 1 | 1 | -0 |
| 5 | 0 | 0 | 25 | 0.79 | 1 | 1 | -0.0001 |
| 5 | 0 | -0.3 | 10 | 0.571 | 1 | 1.019 | 0.0193 |
| 5 | 0 | -0.3 | 16 | 0.692 | 1 | 1.015 | 0.0155 |
| 5 | 0 | -0.3 | 25 | 0.79 | 1 | 1.013 | 0.0127 |
| 5 | 0 | -0.6 | 10 | 0.571 | 1 | 1.063 | 0.0634 |
| 5 | 0 | -0.6 | 16 | 0.692 | 1 | 1.05 | 0.0501 |
| 5 | 0 | -0.6 | 25 | 0.79 | 1 | 1.041 | 0.0407 |
| 5 | 0.262 | 0 | 10 | 0.57 | 2.085 | 2.063 | -0.0106 |
| 5 | 0.262 | 0 | 16 | 0.689 | 2.088 | 2.069 | -0.0093 |
| 5 | 0.262 | 0 | 25 | 0.786 | 2.086 | 2.068 | -0.0088 |
| 5 | 0.262 | -0.3 | 10 | 0.574 | 2.091 | 2.107 | 0.0075 |
| 5 | 0.262 | -0.3 | 16 | 0.699 | 2.093 | 2.108 | 0.0073 |
| 5 | 0.262 | -0.3 | 25 | 0.799 | 2.093 | 2.103 | 0.0049 |
| 5 | 0.262 | -0.6 | 10 | 0.578 | 2.126 | 2.204 | 0.0366 |
| 5 | 0.262 | -0.6 | 16 | 0.71 | 2.125 | 2.185 | 0.0281 |
| 5 | 0.262 | -0.6 | 25 | 0.813 | 2.125 | 2.168 | 0.0201 |
| 5 | 0.531 | 0 | 10 | 0.567 | 4.457 | 4.335 | -0.0275 |
| 5 | 0.531 | 0 | 16 | 0.682 | 4.437 | 4.316 | -0.0275 |
| 5 | 0.531 | 0 | 25 | 0.777 | 4.454 | 4.337 | -0.0266 |
| 5 | 0.531 | -0.3 | 10 | 0.574 | 4.44 | 4.423 | -0.0039 |
| 5 | 0.531 | -0.3 | 16 | 0.7 | 4.43 | 4.417 | -0.0032 |
| 5 | 0.531 | -0.3 | 25 | 0.8 | 4.436 | 4.425 | -0.003 |
| 5 | 0.531 | -0.6 | 10 | 0.584 | 4.55 | 4.656 | 0.0231 |
| 5 | 0.531 | -0.6 | 16 | 0.724 | 4.536 | 4.618 | 0.0176 |
| 5 | 0.531 | -0.6 | 25 | 0.831 | 4.542 | 4.62 | 0.0165 |
| 15 | 0 | 0 | 10 | 0.296 | 1 | 1 | -0.0002 |
| 15 | 0 | 0 | 16 | 0.413 | 1 | 1 | 0.0001 |
| 15 | 0 | 0 | 25 | 0.532 | 1 | 1 | -0.0004 |
| 15 | 0 | -0.3 | 10 | 0.296 | 1 | 1.007 | 0.0072 |
| 15 | 0 | -0.3 | 16 | 0.413 | 1 | 1.007 | 0.0067 |
| 15 | 0 | -0.3 | 25 | 0.532 | 1 | 1.006 | 0.0063 |
| 15 | 0 | -0.6 | 10 | 0.296 | 1 | 1.026 | 0.0259 |
| 15 | 0 | -0.6 | 16 | 0.413 | 1 | 1.025 | 0.0248 |
| 15 | 0 | -0.6 | 25 | 0.532 | 1 | 1.021 | 0.0213 |
| 15 | 0.262 | 0 | 10 | 0.298 | 2.087 | 2.078 | -0.0043 |
| 15 | 0.262 | 0 | 16 | 0.414 | 2.087 | 2.078 | -0.0044 |
| 15 | 0.262 | 0 | 25 | 0.532 | 2.083 | 2.074 | -0.0045 |
| 15 | 0.262 | -0.3 | 10 | 0.288 | 2.091 | 2.096 | 0.0025 |
| 15 | 0.262 | -0.3 | 16 | 0.409 | 2.093 | 2.097 | 0.0023 |
| 15 | 0.262 | -0.3 | 25 | 0.533 | 2.092 | 2.096 | 0.0021 |
| 15 | 0.262 | -0.6 | 10 | 0.278 | 2.128 | 2.155 | 0.013 |
| 15 | 0.262 | -0.6 | 16 | 0.404 | 2.125 | 2.148 | 0.0109 |
| 15 | 0.262 | -0.6 | 25 | 0.536 | 2.127 | 2.143 | 0.0075 |
| 15 | 0.531 | 0 | 10 | 0.306 | 4.453 | 4.399 | -0.0122 |
| 15 | 0.531 | 0 | 16 | 0.418 | 4.446 | 4.384 | -0.0141 |

| base_rate | beta_logRR_perSD | corr_pop_dep | threshold | mean_suppressed | RR_true | RR_cc | RR_relbias |
|---|---|---|---|---|---|---|---|
| 15 | 0.531 | 0 | 25 | 0.53 | 4.436 | 4.365 | -0.0162 |
| 15 | 0.531 | -0.3 | 10 | 0.287 | 4.435 | 4.429 | -0.0013 |
| 15 | 0.531 | -0.3 | 16 | 0.409 | 4.443 | 4.438 | -0.0013 |
| 15 | 0.531 | -0.3 | 25 | 0.534 | 4.428 | 4.418 | -0.0025 |
| 15 | 0.531 | -0.6 | 10 | 0.262 | 4.54 | 4.571 | 0.0067 |
| 15 | 0.531 | -0.6 | 16 | 0.397 | 4.544 | 4.57 | 0.0057 |
| 15 | 0.531 | -0.6 | 25 | 0.538 | 4.551 | 4.576 | 0.0052 |
| 50 | 0 | 0 | 10 | 0.093 | 1 | 1 | 0 |
| 50 | 0 | 0 | 16 | 0.156 | 1 | 1 | 0 |
| 50 | 0 | 0 | 25 | 0.237 | 1 | 1 | 0 |
| 50 | 0 | -0.3 | 10 | 0.093 | 1 | 1.002 | 0.0015 |
| 50 | 0 | -0.3 | 16 | 0.156 | 1 | 1.002 | 0.0018 |
| 50 | 0 | -0.3 | 25 | 0.238 | 1 | 1.002 | 0.0021 |
| 50 | 0 | -0.6 | 10 | 0.093 | 1 | 1.006 | 0.0059 |
| 50 | 0 | -0.6 | 16 | 0.156 | 1 | 1.007 | 0.0069 |
| 50 | 0 | -0.6 | 25 | 0.237 | 1 | 1.008 | 0.0079 |
| 50 | 0.262 | 0 | 10 | 0.096 | 2.084 | 2.082 | -0.0008 |
| 50 | 0.262 | 0 | 16 | 0.159 | 2.087 | 2.084 | -0.0011 |
| 50 | 0.262 | 0 | 25 | 0.241 | 2.086 | 2.083 | -0.0013 |
| 50 | 0.262 | -0.3 | 10 | 0.085 | 2.091 | 2.091 | 0.0004 |
| 50 | 0.262 | -0.3 | 16 | 0.147 | 2.094 | 2.096 | 0.0007 |
| 50 | 0.262 | -0.3 | 25 | 0.229 | 2.091 | 2.093 | 0.0006 |
| 50 | 0.262 | -0.6 | 10 | 0.073 | 2.126 | 2.131 | 0.0024 |
| 50 | 0.262 | -0.6 | 16 | 0.133 | 2.129 | 2.133 | 0.0018 |
| 50 | 0.262 | -0.6 | 25 | 0.215 | 2.126 | 2.129 | 0.0015 |
| 50 | 0.531 | 0 | 10 | 0.106 | 4.46 | 4.448 | -0.0026 |
| 50 | 0.531 | 0 | 16 | 0.17 | 4.44 | 4.421 | -0.0042 |
| 50 | 0.531 | 0 | 25 | 0.25 | 4.445 | 4.42 | -0.0057 |
| 50 | 0.531 | -0.3 | 10 | 0.084 | 4.432 | 4.43 | -0.0004 |
| 50 | 0.531 | -0.3 | 16 | 0.145 | 4.436 | 4.433 | -0.0005 |
| 50 | 0.531 | -0.3 | 25 | 0.228 | 4.435 | 4.434 | -0.0003 |
| 50 | 0.531 | -0.6 | 10 | 0.058 | 4.555 | 4.56 | 0.001 |
| 50 | 0.531 | -0.6 | 16 | 0.115 | 4.544 | 4.55 | 0.0013 |
| 50 | 0.531 | -0.6 | 25 | 0.197 | 4.549 | 4.553 | 0.0009 |
| 150 | 0 | 0 | 10 | 0.021 | 1 | 1 | -0 |
| 150 | 0 | 0 | 16 | 0.042 | 1 | 1 | 0.0001 |
| 150 | 0 | 0 | 25 | 0.075 | 1 | 1 | -0.0002 |
| 150 | 0 | -0.3 | 10 | 0.021 | 1 | 1 | 0 |
| 150 | 0 | -0.3 | 16 | 0.042 | 1 | 1 | 0.0003 |
| 150 | 0 | -0.3 | 25 | 0.075 | 1 | 1.001 | 0.0005 |
| 150 | 0 | -0.6 | 10 | 0.021 | 1 | 1.001 | 0.0007 |
| 150 | 0 | -0.6 | 16 | 0.041 | 1 | 1.001 | 0.0013 |
| 150 | 0 | -0.6 | 25 | 0.075 | 1 | 1.002 | 0.0019 |
| 150 | 0.262 | 0 | 10 | 0.022 | 2.086 | 2.085 | -0.0002 |
| 150 | 0.262 | 0 | 16 | 0.044 | 2.082 | 2.081 | -0.0002 |
| 150 | 0.262 | 0 | 25 | 0.078 | 2.087 | 2.087 | -0.0004 |
| 150 | 0.262 | -0.3 | 10 | 0.017 | 2.092 | 2.093 | 0.0001 |
| 150 | 0.262 | -0.3 | 16 | 0.036 | 2.092 | 2.092 | 0.0003 |
| 150 | 0.262 | -0.3 | 25 | 0.067 | 2.094 | 2.094 | 0.0001 |
| 150 | 0.262 | -0.6 | 10 | 0.013 | 2.128 | 2.128 | 0.0002 |
| 150 | 0.262 | -0.6 | 16 | 0.028 | 2.126 | 2.127 | 0.0002 |
| 150 | 0.262 | -0.6 | 25 | 0.056 | 2.126 | 2.126 | 0.0001 |
| 150 | 0.531 | 0 | 10 | 0.027 | 4.448 | 4.446 | -0.0004 |
| 150 | 0.531 | 0 | 16 | 0.05 | 4.457 | 4.454 | -0.0008 |
| 150 | 0.531 | 0 | 25 | 0.087 | 4.448 | 4.442 | -0.0013 |
| 150 | 0.531 | -0.3 | 10 | 0.016 | 4.429 | 4.428 | -0.0001 |
| 150 | 0.531 | -0.3 | 16 | 0.035 | 4.434 | 4.434 | -0.0001 |

| base_rate | beta_logRR_perSD | corr_pop_dep | threshold | mean_suppressed | RR_true | RR_cc | RR_relbias |
|---|---|---|---|---|---|---|---|
| 150 | 0.531 | -0.3 | 25 | 0.066 | 4.436 | 4.436 | -0 |
| 150 | 0.531 | -0.6 | 10 | 0.008 | 4.554 | 4.555 | 0.0002 |
| 150 | 0.531 | -0.6 | 16 | 0.02 | 4.552 | 4.552 | 0.0001 |
| 150 | 0.531 | -0.6 | 25 | 0.043 | 4.553 | 4.554 | 0.0001 |

### Supplementary Table S2. Calibrated sensitivity (corr = -0.30)

Population-weighted vs unweighted; Poisson (overdisp 0) and overdispersed (0.5).

| base_rate | corr_pop_dep | threshold | overdisp | mean_suppressed | RR_true | RR_cc_wt | RR_cc_unwt | relbias_wt | relbias_unwt |
|---|---|---|---|---|---|---|---|---|---|
| 5 | -0.3 | 16 | 0 | 0.7 | 4.425 | 4.413 | 4.424 | -0.003 | -0 |
| 5 | -0.3 | 16 | 0.5 | 0.731 | 4.437 | 4.381 | 4.351 | -0.013 | -0.019 |
| 15 | -0.3 | 16 | 0 | 0.409 | 4.433 | 4.425 | 4.423 | -0.002 | -0.002 |
| 15 | -0.3 | 16 | 0.5 | 0.479 | 4.439 | 4.406 | 4.336 | -0.007 | -0.023 |
| 50 | -0.3 | 16 | 0 | 0.145 | 4.43 | 4.429 | 4.434 | -0 | 0.001 |
| 50 | -0.3 | 16 | 0.5 | 0.22 | 4.444 | 4.458 | 4.371 | 0.003 | -0.016 |
| 150 | -0.3 | 16 | 0 | 0.035 | 4.442 | 4.442 | 4.439 | 0 | -0.001 |
| 150 | -0.3 | 16 | 0.5 | 0.079 | 4.437 | 4.477 | 4.401 | 0.009 | -0.008 |

### Supplementary Table S3. STROBE checklist (cross-sectional)

| STROBE item | Recommendation | Location |
|---|---|---|
| 1 Title/abstract | Design in title; structured abstract | Title; Abstract |
| 2 Background | Scientific background, rationale | Introduction |
| 3 Objectives | Stated objective/aim | Introduction (aim) |
| 4 Study design | Key design elements | Methods 1 |
| 5 Setting | Setting, locations, dates | Methods 1,4 |
| 6 Participants | Units, eligibility | Methods 2 |
| 7 Variables | Exposure, outcome, covariates | Methods 3-4 |
| 8 Data sources/measurement | Sources, measurement | Methods 1,3,4 |
| 9 Bias | Sources of bias addressed | Methods 6; Discussion |
| 10 Study size | How arrived at | Methods 2,7 |
| 11 Quantitative variables | Handling, groupings | Methods 3,5 |
| 12 Statistical methods | Methods, subgroups, sensitivity | Methods 5-9 |
| 13 Participants (results) | Numbers at each stage | Results (sample) |
| 14 Descriptive data | Characteristics | Table 1 |
| 15 Outcome data | Outcomes/events | Tables 2-3 |
| 16 Main results | Estimates + CIs | Tables 2; Figs 1,3 |
| 17 Other analyses | Subgroups/sensitivity/sim | Methods 7-9; Fig 3 |
| 18 Key results | Summary re objectives | Discussion 1 |
| 19 Limitations | Limitations, bias direction | Discussion 5 |
| 20 Interpretation | Cautious overall interpretation | Discussion 2-3,7 |
| 21 Generalisability | External validity | Discussion 2,6 |
| 22 Funding | Funding role | Statements |

Simulation reporting follows ADEMP (Morris et al., 2019): aims, data-generating mechanisms, estimands, methods, and performance measures (bias, percent bias, Monte Carlo SE) are specified in Methods section 7.
